# Effects of vitamin D deficiency on blood lipids and bone metabolism: a large cross-sectional study

**DOI:** 10.1101/2022.04.12.22273773

**Authors:** Peng Gu, Bin Pu, BaiHang Chen, XiaoHui Zheng, ZhanPeng Zeng, WeiDong Luo

## Abstract

**Purpose:** To investigate the relationship between serum HDL-C and spinal bone mineral density (BMD) under different serum 25-hydroxyvitamin D (25 (OH) D) levels in adults over 40 years old and to explore its mechanism.

**Methods:** Participants over the age of 40 with data on HDL-C, 25 (OH) D, spinal BMD, and other variables in NHANES 2007–2010 were included in the final analysis. A weighted multiple linear regression model was used to evaluate the association between serum HDL-C and spinal BMD in different gender, ages and serum 25 (OH) D levels.

**Results:** A total of 3599 subjects aged ≥ 40 years old were included in this study. Univariate analysis of the complete correction model showed a negative correlation between serum HDL-C and spinal BMD. In the two subgroups of serum 25(OH)D, we found that the higher the serum HDL-C in the female with serum 25 (OH) D < 75nmol/L aged 40-59 years old, the lower the total spinal BMD, and a similar relationship was found in the lumbar spine (L1-L4). However, no similar relationship was found in all populations with serum 25 (OH) D ≥ 75nmol/L and males with serum 25 (OH) D < 75nmol/L.

**Conclusion:** Among Americans over the age of 40, the increase of serum HDL-C is related to decreased BMD of spine only in women aged 40-59 years with vitamin D insufficiency or deficiency.

**Author summary:** We performed a cross-sectional study using the National Health Examination and Nutrition Survey (NHANES) data. We found that increased serum high-density lipoprotein cholesterol (HDL-C) during vitamin D deficiency is a potential risk factor for osteopenia or osteoporosis in middle-aged and elderly American women.

## Introduction

Osteoporosis (OP) is a systemic skeletal disease common in the elderly, which is characterized by reduced bone mass per unit volume, decreased bone strength, destruction of bone microstructure and increased risk of fracture(1). OP is often associated with brittle fractures, with approximately 1/2 of women and 1/5 of men expected to experience an osteoporotic fracture during their lifetime(2). BMD is a standard parameter to evaluate bone health. There are many risk factors for BMD reduction, including genetic, lifestyle and nutrition factors, which have been identified to be related to BMD reduction(3-5). The existing evidence also showed that abnormal blood lipid metabolism(6-8) and serum vitamin D deficiency(9, 10) were closely related to osteoporosis. Patients with hyperlipidemia can have bone loss and osteoporotic fracture simultaneously. Patients with OP and osteopenia are also often complicated with abnormal blood lipid metabolism(11), especially the abnormal metabolism of serum HDL-C(12). Serum vitamin D is an essential factor in regulating the balance of calcium, phosphorus and bone metabolism, and the appropriate nutritional status of vitamin D is significant for the maintenance of bone health(13). Vitamin D deficiency is associated with various diseases harmful to bone health, including diabetes, chronic obstructive pulmonary disease(14), a variety of autoimmune diseases(15) and dyslipidemia(16).

Existing studies indicated that serum HDL-C levels were negatively correlated with bone status in postmenopausal women with vitamin D deficiency(17). However, this relationship was only evaluated in postmenopausal women and did not extend to all age groups and men, and it was a single-center clinical study with a small sample size. This study conducted a cross-sectional study combining data from two cycles (2007-2008 and 2009-2010) of the National Health and Nutrition Examination Survey (NHANES) to provide more evidence-based references further. It was objective for the study to evaluate the association between serum HDL-C and BMD at different serum vitamin D levels and to provide guidance for regulating serum vitamin D and HDL-C levels to reduce the risk of osteopenia or OP.

## Methods

### Data source and study design

NHANES is a series of continuous cross-sectional surveys conducted jointly by the National Center For Health Statistics and the Centers for Disease Control And Prevention to include information about a representative general non-institutionalized population of all ages in the United States to assess their nutrition and health status. It is reviewed and approved by the NCHS Research Ethics Review Board and obtained all participants’ written and informed consent. This survey includes two parts: interview and physical examination data. The interview part mainly collects demographic data, questionnaire data, and dietary data. The physical examination mainly includes massive laboratory data and examination data. Each individual in the database is replaced with a unique ID and does not include any personally identifiable private information.

This study combines the data of NHANES 2007-2010. Inclusion criteria included: (1) participants ≥ 40 years old, (2) participants with available spinal BMD, HDL-C, and 25 (OH) D data. Exclusion criteria included: (1) subjects who have been treated for osteoporosis (who have treated osteoporosis); (2) prednisone or cortisone every day (prednisone or cortisone tablets almost every day for a month or more?); (3) subjects with kidney weakness or failure (have you been told by your doctor or other health professionals that you have kidney weakness or failure? Does not include kidney stones, bladder infection or incontinence.)(4) subjects with missing data of other variables were excluded. Finally, among 20686 participants, through strict eligibility criteria, a total of 3599 participants were included in the study. (Fig 1)Data collection/analysis for this study was conducted from January to March 2022. The date that participants were recruited to the study and the date range in which human subjects’ data/samples visit https://wwwn.cdc.gov/nchs/nhanes/Default.aspx.

**Figure1.**
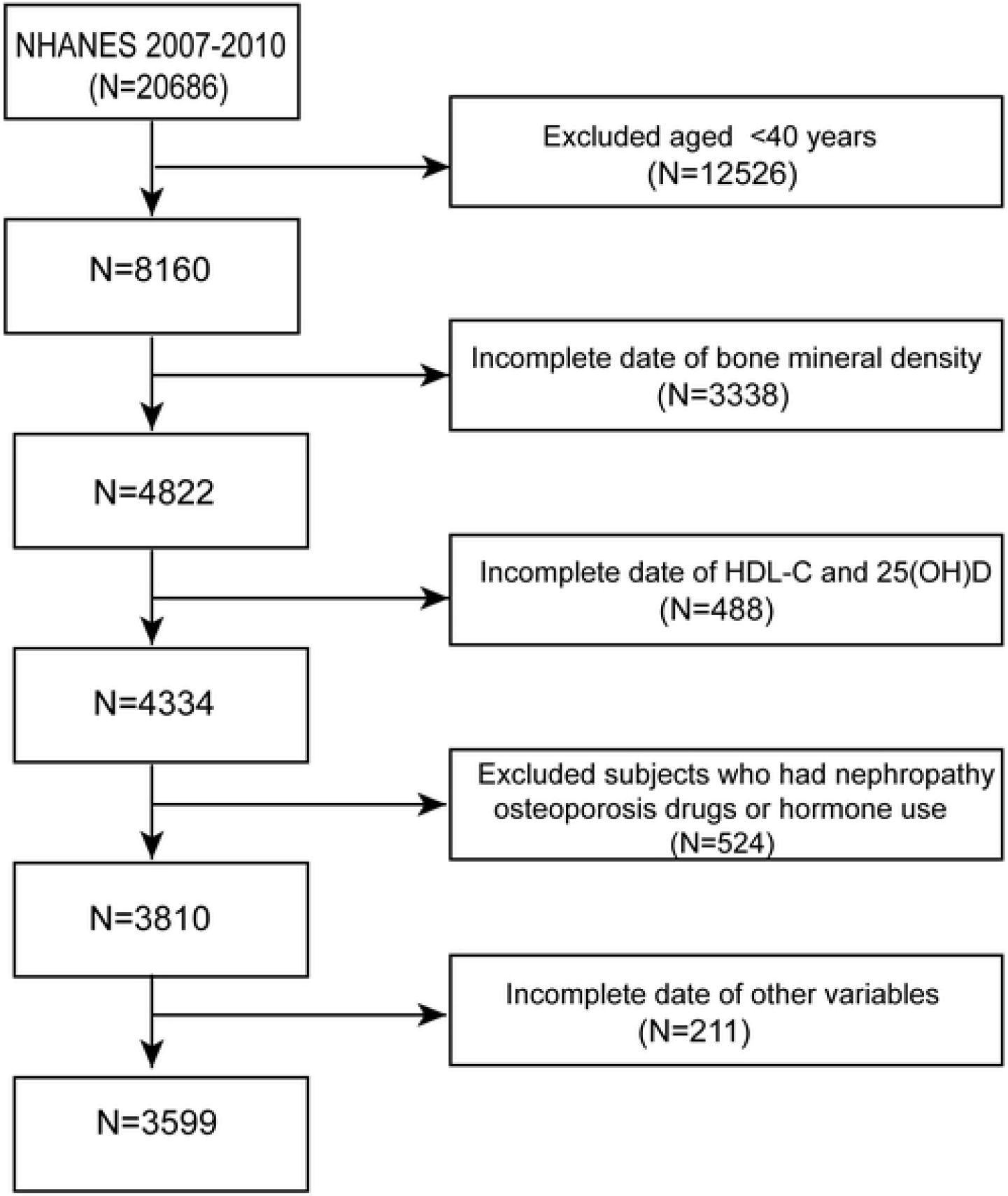

### Measurement of serum 25(OH)D level

For the two cycles of NHANES 2007-2010, 25-hydroxyvitamin D3 (25(OH)D3), epi-25-hydroxyvitamin D3 (epi-25(OH)D3) and 25-hydroxyvitamin D2 (25(OH)D2) were determined by ultra-high-performance liquid chromatography-tandem mass spectrometry. Total 25-hydroxyvitamin D is the sum of 25-hydroxyvitamin D2 and 25-hydroxyvitamin D3 but does not include table 25-hydroxyvitamin D3. Detailed instructions on specimen collection and processing can be found on the NHANES websites(https://wwwn.cdc.gov/nchs/nhanes/2007-2008/VID_E.htm and https://wwwn.cdc.gov/nchs/nhanes/2009-2010/VID_F.htm). According to the guidelines for the evaluation, treatment, and prevention of vitamin D deficiency issued by the American Endocrine Association in July 2014, 25 (OH) D levels were divided into two groups: serum vitamin D insufficiency or deficiency group with total 25 (OH) D concentration < 75nmol/L and serum vitamin D normal group with total 25 (OH) D concentration ≥ 75nmol/L(18).

### Measurement of serum HDL-C level

Subjects’ sera were collected to measure HDL-C levels and all samples were analyzed at the University of Minnesota Lipid Laboratory using a Roche modular P chemical analyzer. Magnesium sulfate/glucose solution, cholesterol oxidase and polyethylene glycol coupled cholesterol esterase and other reagents were used for determination. These reagents are related to a laboratory method reference material in the cholesterol reference method laboratory network certified by the Centers for Disease Control and Prevention (CDC) Lipid Standardization Program, with no differences in the generation assays between measurements.

### Measurement of results and covariates

The outcome variables are the spinal BMD (total spinal BMD, L1-L4BMD) measured by dual-energy X-ray absorptiometry on the Hologic Discovery model A densitometer (Hologic, Inc., Bedford, Massachusetts). All scans were analyzed using Hologic APEX3.0 software. Covariates include demographic data, such as age, gender, race, education level, marital status, body mass index (kg/m^2^), C-reactive protein (mmol/L), serum total calcium concentration (mmol/L), phosphorus (mmol/L), total cholesterol (mmol/L), alkaline phosphatase (U/L), aspartate aminotransferase (U/L), glutamic pyruvic transaminase (U/L), smoking status, alcohol consumption, hypertension, diabetes. Covariates were collected through family interviews, physical examination, laboratory measurements, and questionnaires. For more details on data collection, visit https://wwwn.cdc.gov/nchs/nhanes/ContinuousNhanes/Default.aspx?BeginYear=2007 and https://wwwn.cdc.gov/nchs/nhanes/ContinuousNhanes/Default.aspx?BeginYear=2009.

### Statistical analysis

To account for oversampling in complex survey design, survey nonresponse, and poststratification, the 2007/2008 and 2009/2010 cycles were combined, and 4-y sampling weights were constructed by using one-two of the 2-y sampling weight (WTMEC2YR) constructed by the NHANES.The baseline characteristics of all study participants were described by average (continuous variables) or proportions (classified variables). Weighted multiple linear regression model evaluated the linear relationship between serum HDL-C and BMD. Unadjusted and multivariate-adjusted logical regression analyses were used to calculate the P value and the corresponding 95% confidence interval (CI) to determine the relationship between serum HDL-C levels and spinal BMD. The coarse model was adjusted to no variable. The multivariate model included age, sex, race, smoking status, alcohol consumption, hypertension, diabetes, BMI, CRP, Ca, P, TC, ALP, AST, ALT. We selected these confounding factors based on their association with more than 10% changes in the estimated results or effects of interest. Multiple linear regression models were used for subgroup analysis, and the linear relationship between serum HDL-C and spinal BMD in different populations was evaluated by sex, age, and serum 25 (OH) D stratification. P < 0.05 is defined as significant. Multi-factor histogram was used to describe the independent association between BMD and HDL-C in different parts of a specific population. All analyses were conducted using R (version 4.0.3) and EmpowerStats software (http://www.empowerstats.com). The figures were generated using GraphPad Prism 9.0.0(121) (https://www.graphpad.com/).

## Results

### Participant selection and baseline characteristics

Participants’ information was extracted from the NHANES database 2007-2010. (i) exclusion of subjects younger than 40 (n = 12526), (ii) exclusion of subjects without spinal BMD data (n = 3338), (iii) exclusion of subjects without HDL-C, 25 (OH) D data (n = 488) (iv) excluding subjects who had treated osteoporosis, used or continued hormone therapy, and suffered from kidney weakness or failure (n=524) (v) excluded the missing values of other variables (n = 211). After that, 3599 participants (Fig 1) participated in the final analysis. The database includes 1814 men and 1785 women, the average age of men was 54.09 ±10.33 years old, most of them were non-Hispanic white (74.64%), college degree or above (58.47%), smoking (47.84%), drinking (at least 12 drinks in the past year) (85.03%), high blood pressure (35.98%), diabetes (10.23%). Male BMI (28.53 ±4.70kg/m^2^), 25 (OH) D (67.04 ±22.53nmol/l), ALT (30.77 ±25.05U/L), AST (28.49 ±16.87U), ALP (66.98 ±20.58U/L), TC (5.19 ±1.09mmol/L), Ca (2.35 ±0.09mmol/L), P (1.16 ±0.18mmol/L), CRP (0.28 ±0.59mmol/L). The average BMD of total spine, L1, L2, L3 and L4 was 1.06 ±0.15g/cm^2^, 1.00 ±0.14g/cm^2^, 1.07 ±0.15g/cm^2^, 1.08 ±0.16g/cm^2^ and 1.08 ±0.16g/cm^2^, respectively. The average age of women was 54.11 ±10.44. most of the women were non-Hispanic whites (74.27%), college degree or above (58.96%), smoking (58.92%), alcohol consumption (67.82%), hypertension (33.74%), diabetes (8.69%), BMI (28.12 ±6.26kg/m^2^). 25 (OH) D (70.99 ±27.31nmol/l), ALT (22.24 ±13.05U/L), AST (24.51 ±11.28U/L), ALP (68.47 ±22.09U/L), TG (5.39 ±1.04mmol/L), Ca (1.25 ±0.17mmol/L), P (2.36 ±0.09mmol/L), CRP (0.41 ±0.76mmol/L). The average BMD of total spine, L1, L2, L3 and L4 was 1.00 ±0.14g/cm^2^, 0.90 ±0.15g/cm^2^, 1.00 ±0.15g/cm^2^,1.04±0.15g/cm^2^,1.04 ±0.15g/cm^2^, 1.04 ±0.15g/cm^2^, respectively (Table 1).

**Table 1.** Characteristics of study sample: US population aged ≥ 40 years in the NHANES 2007-2010.

| Characteristics | Total number of participants (n=3599) |  | P |
| --- | --- | --- | --- |
|  | Men (n=1814) | Women (n=1785) |  |
| Age (years, mean ± SD) | 54.09 ± 10.33 | 54.11 ± 10.44 | 0.9449 |
| Race/ethnicity, n (%) |  |  | 0.6246 |
| Mexican American | 338 (7.32) % | 362 (7.08) % |  |
| Other Hispanic | 194 (3.99) % | 201 (4.02) % |  |
| Non-Hispanic White | 893 (74.64) % | 848 (74.27) % |  |
| Non-Hispanic Black | 321 (8.78) % | 317 (10.07) % |  |
| Other Race | 68 (5.27) % | 57 (4.56) % |  |
| Education, n (%) |  |  | 0.3241 |
| Under high school | 561 (18.27) % | 533 (17.47) % |  |
| High school or equivalent | 415 (23.26) % | 403 (23.57) % |  |
| Some College or AA degree | 427 (26.88) % | 484 (29.35) % |  |
| College Graduate or above | 411 (31.59) % | 365 (29.61) % |  |
| BMI, mean ± SD (kg/m<sup>2</sup>) | 28.53 ± 4.70 | 28.12 ± 6.26 | 0.0286 |
| Smoking, n(%) |  |  | <0.0001 |
| Never | 441 (22.96) % | 318 (18.19) % |  |
| Former | 584 (29.19) % | 381 (22.90) % |  |
| Current | 789 (47.84) % | 1086 (58.92) % |  |
| Had at least 12 alcohol drinks past one year? n (%) |  |  | <0.0001 |
| Yes | 1501 (85.03) % | 1058 (67.82) % |  |
| No | 313 (14.97) % | 727 (32.18) % |  |
| Marital status |  |  | <0.0001 |
| Live with someone | 1327 (77.20) % | 1043 (66.13) % |  |
| Live alone | 487 (22.80) % | 742 (33.87) % |  |
| Hypertension, n (%) |  |  | 0.1586 |
| Yes | 714 (35.98) % | 715 (33.74) % |  |
| No | 1100 (64.02) % | 1070 (66.26) % |  |
| Diabetes, n (%) |  |  | 0.2750 |
| Yes | 254 (10.23) % | 247 (8.69) % |  |
| No | 1521 (87.97) % | 1505 (89.61) % |  |
| Borderline | 39 (1.79) % | 33 (1.70) % |  |
| Total 25(OH)D (nmol/l) | 67.04 ± 22.53 | 70.99 ± 27.31 | <0.0001 |
| Phosphorus (mmol/L, mean ± SD) | 1.16 ± 0.18 | 1.25 ± 0.17 | <0.0001 |
| Total calcium (mmol/L, mean ± SD) | 2.35 ± 0.09 | 2.36 ± 0.09 | 0.1269 |
| CRP (mmol/L, mean ± SD) | 0.28 ± 0.59 | 0.41 ± 0.76 | <0.0001 |
| HDL-C (mmol/L, mean ± SD) | 1.24 ± 0.38 | 1.55 ± 0.45 | <0.0001 |
| TC (mmol/L, mean ± SD) | 5.19 ± 1.09 | 5.39 ± 1.04 | <0.0001 |
| ALP (U/L, mean ± SD) | 66.98 ± 20.58 | 68.47 ± 22.09 | 0.0359 |
| ALT (U/L, mean ± SD) | 30.77 ± 25.05 | 22.24 ± 13.05 | <0.0001 |
| AST (U/L, mean ± SD) | 28.49 ± 16.87 | 24.51 ± 11.28 | <0.0001 |
| Total spine BMD (g/cm<sup>2</sup>, mean ± SD) | 1.06 ± 0.15 | 1.00 ± 0.14 | <0.0001 |
| L1 BMD (g/cm<sup>2</sup>, mean ± SD) | 1.00 ± 0.14 | 0.90 ± 0.15 | <0.0001 |
| L2 BMD (g/cm<sup>2</sup>, mean ± SD) | 1.07 ± 0.15 | 1.00 ± 0.15 | <0.0001 |
| L3 BMD (g/cm<sup>2</sup>, mean ± SD) | 1.08 ± 0.16 | 1.04 ± 0.15 | <0.0001 |
| L4 BMD (g/cm<sup>2</sup>, mean ± SD) | 1.08 ± 0.16 | 1.04 ± 0.15 | <0.0001 |

NHANES = National Health and Nutrition Examination Survey; SD = standard deviation; Values are mean ± SD or n (%).
HDL-C = high-density lipoprotein cholesterol; BMI = Body mass index; CRP = C-reactive protein; TC = Cholesterol; ALP = Alkaline phosphatase; ALT = Alanine aminotransferase; AST = Aspartate aminotransferase; BMD = bone mineral density ; CI = confidence interval; $\beta$ regression coefficient

### Relationship between serum HDL-C and BMD

Serum HDL-C was negatively correlated with BMD, model 1(without adjusting covariates). After adjusting for confounding factors, model 2 (age, sex, race/ethnicity were adjusted), and model 3 (all covariates were adjusted), all have similar results. There were significant differences in trend test results between the two groups (P < 0.05) (Table 2).

**Table 2.** Correlation between high-density lipoprotein-cholesterol and bone mineral density

| BMD | Model1 |  | Model2 |  | Model3 |  |
| --- | --- | --- | --- | --- | --- | --- |
| | $\beta$(95%CI) | P | $\beta$(95%CI) | P | $\beta$(95%CI) | P |
| Total spine | -0.055 (-0.066, -0.044) | <0.00001 | -0.043 (-0.054, -0.032) | <0.00001 | -0.018 (-0.031, -0.006) | 0.00373 |
| L1 | -0.082 (-0.093, -0.071) | <0.00001 | -0.053 (-0.064, -0.042) | <0.00001 | -0.026 (-0.038, -0.013) | 0.00005 |
| L2 | -0.062 (-0.073, -0.050) | <0.00001 | -0.045 (-0.056, -0.033) | <0.00001 | -0.021 (-0.034, -0.008) | 0.00133 |
| L3 | -0.044 (-0.055, -0.032) | <0.00001 | -0.041 (-0.053, -0.028) | <0.00001 | -0.016 (-0.029, -0.002) | 0.02179 |
| L4 | -0.041 (-0.053, -0.030) | <0.00001 | -0.037 (-0.049, -0.025) | <0.00001 | -0.014 (-0.027, -0.000) | 0.04453 |

Model1, no covariates were adjusted
Model2, age,sex,race / ethnicity, were adjusted

### Relationship between serum HDL-C and BMD at different levels of serum 25 (OH) D

In all covariant adjusted models, we observed that when serum 25 (OH) D < 75nmol/l, serum HDL-C was negatively correlated with total spine and L1-L4 BMD (P<0.05). However, when serum 25 (OH) D ≥ 75nmol/l, there was no correlation between serum HDL-C and total spine and L1-L4 BMD (P> 0.05) (Table 3).

**Table 3.** Correlation between high-density lipoprotein cholesterol and bone mineral density based on 25(OH)D status classification

| BMD | Total 25(OH)<75 (nmol/l) (n=2565) |  | Total 25(OH)≥75 (nmol/l) (n=1034) |  |
| --- | --- | --- | --- | --- |
|  | Adjust β (95% CI) | P | Adjust β (95% CI) | P |
| Total spine | -0.024 (-0.040, -0.008) | 0.0033 | -0.007 (-0.028, 0.014) | 0.5062 |
| L1 | -0.032 (-0.048, -0.016) | <0.0001 | -0.013 (-0.033, 0.008) | 0.2315 |
| L2 | -0.025 (-0.042, -0.009) | 0.0025 | -0.010 (-0.032, 0.012) | 0.3622 |
| L3 | -0.020 (-0.037, -0.002) | 0.0289 | -0.008 (-0.031, 0.015) | 0.4913 |
| L4 | -0.022 (-0.039, -0.005) | 0.0131 | -0.000 (-0.023, 0.023) | 0.9929 |

Furthermore, the subgroup analysis of people with serum 25 (OH) D < 75nmol/l by sex and age showed that when serum 25 (OH) D < 75nmol/l, there were differences in the association between serum HDL-C level and BMD in different sex and age groups. When serum 25 (OH) D < 75nmol/l, the spine BMD(total spine and L1-L4) of men of all ages were not correlated with HDL-C (P > 0.05). The total spine, L1 and L4 BMD of female of 40 ≤ aged < 60, the L2 BMD of 40 ≤ Aged < 50 female and the L3 BMD of 50 ≤ Aged < 60 female were negatively correlated with serum HDL-C, while the L2 BMD of 50 ≤ Aged < 60 female, the L3 BMD of 40 ≤ Aged < 50 female and the remaining 60-80 age group did not correlate with serum HDL-C (Table 4). Classification of Women aged 40-59 years with serum 25(OH)D<75nmol/l into low, middle and high groups based on the third quartile interval of serum HDL-C concentration, the results were a negative correlation between different serum HDL-C groups and BMD, and the association between BMD in different lumbar vertebrae was L1 < L2 < L3 < L4 (Fig 2).

**Table 4.** Correlation between high-density lipoprotein cholesterol and bone mineral density based on Gender and Age status classification

| BMD | Age | Male (n=1341) |  | Female (n=1224) |  |
| --- | --- | --- | --- | --- | --- |
| | | Adjust $\beta$ (95% CI) | P | Adjust $\beta$ (95% CI) | P |
| Total spine | 40≤Aged<50 | -0.007 (-0.048, 0.034) | 0.7390 | -0.038 (-0.068, -0.007) | 0.0161 |
| | 50≤Aged<60 | -0.024 (-0.074, 0.026) | 0.3507 | -0.053 (-0.097, -0.010) | 0.0176 |
| | 60≤Aged<70 | 0.008 (-0.045, 0.060) | 0.7749 | -0.015 (-0.060, 0.031) | 0.5271 |
| | 70≤Aged | -0.006 (-0.076, 0.065) | 0.8730 | -0.028 (-0.108, 0.052) | 0.4989 |
| L1 | 40≤Aged<50 | -0.003 (-0.045, 0.039) | 0.8824 | -0.056 (-0.089, -0.024) | 0.0007 |
| | 50≤Aged<60 | -0.036 (-0.085, 0.013) | 0.1538 | -0.063 (-0.110, -0.017) | 0.0076 |
| | 60≤Aged<70 | -0.017 (-0.068, 0.034) | 0.5142 | -0.036 (-0.078, 0.006) | 0.0984 |
| | 70≤Aged | -0.004 (-0.070, 0.062) | 0.9061 | -0.041 (-0.116, 0.034) | 0.2909 |
| L2 | 40≤Aged<50 | -0.016 (-0.058, 0.026) | 0.4512 | -0.033 (-0.066, -0.001) | 0.0448 |
| | 50≤Aged<60 | -0.032 (-0.083, 0.019) | 0.2233 | -0.042 (-0.088, 0.005) | 0.0786 |
| | 60≤Aged<70 | 0.021 (-0.032, 0.074) | 0.4453 | -0.016 (-0.063, 0.030) | 0.4904 |
| | 70≤Aged | -0.013 (-0.085, 0.059) | 0.7291 | -0.052 (-0.130, 0.027) | 0.1994 |
| L3 | 40≤Aged<50 | 0.005 (-0.040, 0.050) | 0.8314 | -0.027 (-0.060, 0.007) | 0.1185 |
| | 50≤Aged<60 | -0.006 (-0.060, 0.049) | 0.8375 | -0.059 (-0.107, -0.012) | 0.0144 |
| | 60≤Aged<70 | 0.020 (-0.038, 0.079) | 0.4962 | -0.013 (-0.064, 0.039) | 0.6255 |
| | 70≤Aged | -0.013 (-0.089, 0.063) | 0.7417 | -0.036 (-0.124, 0.053) | 0.4286 |
| L4 | 40≤Aged<50 | -0.014 (-0.058, 0.030) | 0.5257 | -0.037 (-0.070, -0.005) | 0.0229 |
| | 50≤Aged<60 | -0.028 (-0.083, 0.027) | 0.3198 | -0.048 (-0.094, -0.003) | 0.0396 |
| | 60≤Aged<70 | 0.005 (-0.054, 0.063) | 0.8727 | -0.002 (-0.054, 0.049) | 0.9323 |
| | 70≤Aged | 0.005 (-0.076, 0.086) | 0.9081 | 0.004 (-0.084, 0.092) | 0.9277 |

**Figure2.**
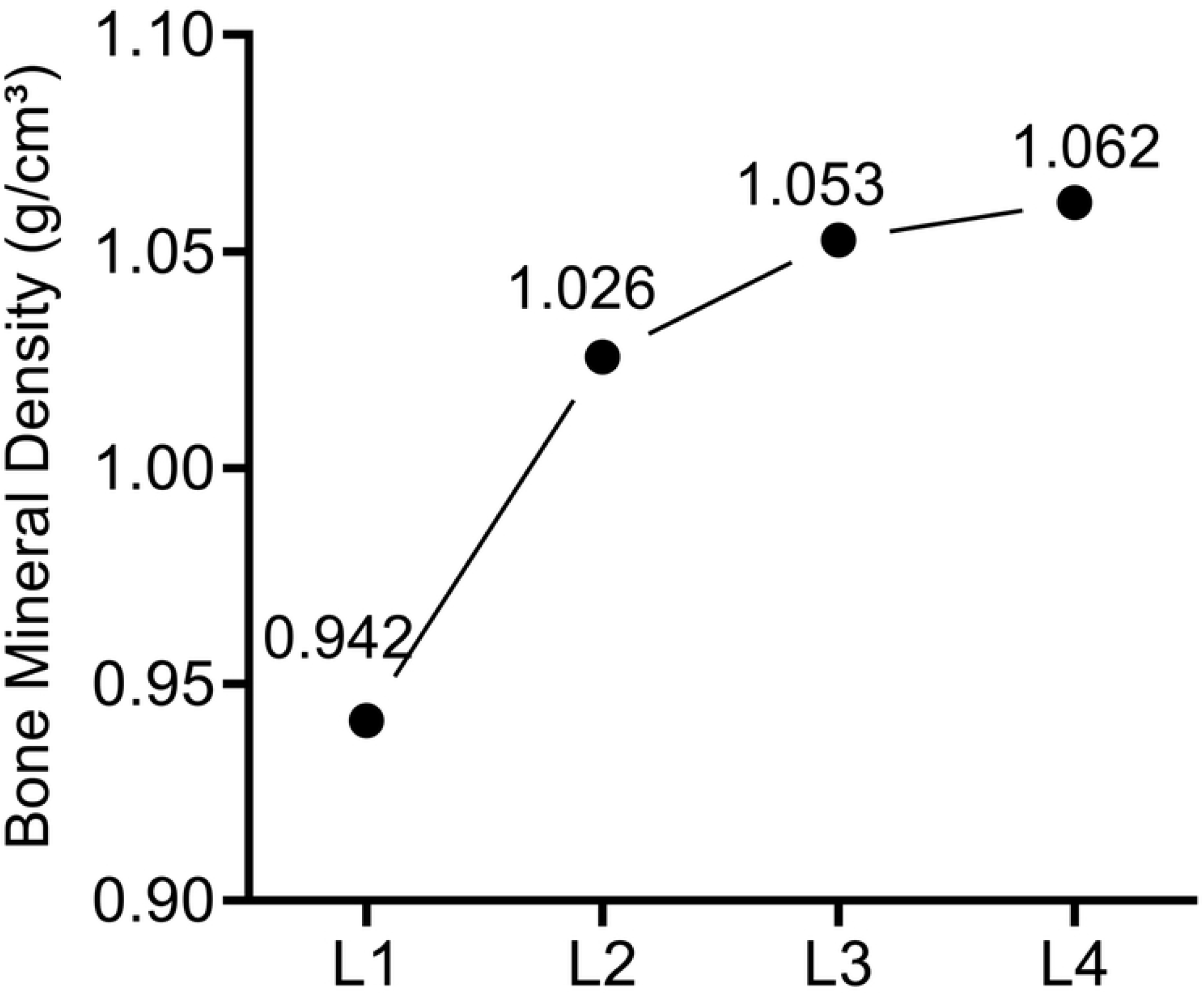

### Relationship between serum HDL-C and BMD based on horizontal stratification of covariates

After stratified analysis of the relationship between serum HDL-C and spinal BMD by using all covariates (the covariates with continuous data as classified variables) as line stratified variables, it was found that the serum HDL-C of non-Hispanic blacks, people living with someone, smokers, people with education below high school level and people with highest ALP quartile had a negative correlation with spinal BMD (Table 5).

**Table 5.** Correlation between high-density lipoprotein cholesterol and bone mineral density based on covariate status classification.

|  |  | TOTAL.SPINE | L1 | L2 | L3 | L4 |
| --- | --- | --- | --- | --- | --- | --- |
| Covariant |  | β(95%CI) | β(95%CI) | β(95%CI) | β(95%CI) | β(95%CI) |
|  |  | P | P | P | P | P |
| RACE | Mexican American | -0.010 (-0.040, 0.020)<br>0.5094 | -0.020 (-0.051, 0.010)<br>0.1877 | -0.006 (-0.037, 0.025)<br>0.7073 | -0.005(-0.037,0.028)<br>0.7672 | -0.011 (-0.043, 0.022)<br>0.5212 |
|  | Other Hispanic | -0.016 (-0.060, 0.027)<br>0.4570 | -0.041 (-0.086, 0.003)<br>0.0692 | -0.013 (-0.057, 0.032)<br>0.5779 | -0.017 (-0.064, 0.029)<br>0.4593 | -0.001 (-0.049, 0.046)<br>0.9517 |
|  | Non-Hispanic White | -0.017 (-0.035, 0.001)<br>0.0720 | -0.021 (-0.039, -0.003)<br>0.0234 | -0.017 (-0.036, 0.002)<br>0.0760 | -0.016 (-0.036, 0.004)<br>0.1111 | -0.014 (-0.034, 0.005)<br>0.1548 |
|  | Non-Hispanic Black | -0.041 (-0.069, -0.013)<br>0.0042 | -0.058 (-0.086, -0.030)<br><0.0001 | -0.052 (-0.081, -0.023)<br>0.0004 | -0.035 (-0.065, -0.005)<br>0.0232 | -0.026 (-0.057, 0.004)<br>0.0865 |
|  | Other Race | 0.001 (-0.079, 0.080)<br>0.9881 | -0.003 (-0.084, 0.078)<br>0.9396 | -0.016 (-0.098, 0.067)<br>0.7084 | 0.009 (-0.079, 0.096)<br>0.8487 | 0.007 (-0.077, 0.092)<br>0.8704 |
| MARITAL | Live with someone | -0.020 (-0.036, -0.005)<br>0.0102 | -0.025 (-0.040, -0.009)<br>0.0016 | -0.021 (-0.037, -0.005)<br>0.0096 | -0.018 (-0.035, -0.001)<br>0.0350 | -0.019 (-0.036, -0.002)<br>0.0287 |
|  | Live alone | -0.016 (-0.037, 0.005)<br>0.1323 | -0.027 (-0.048, -0.005)<br>0.0145 | -0.021 (-0.043, 0.001)<br>0.0571 | -0.014 (-0.037, 0.009)<br>0.2239 | -0.007 (-0.030, 0.015)<br>0.5284 |
| EDUCATION | Under high school | -0.036 (-0.059, -0.013)<br>0.0024 | -0.033 (-0.057, -0.010)<br>0.0050 | -0.035 (-0.058, -0.011)<br>0.0041 | -0.035 (-0.060, -0.009)<br>0.0075 | -0.041 (-0.066, -0.016)<br>0.0014 |
|  | High school or equivalent | -0.023 (-0.050, 0.004)<br>0.0981 | -0.040 (-0.066, -0.014)<br>0.0029 | -0.030 (-0.058, -0.002)<br>0.0381 | -0.013 (-0.043, 0.017)<br>0.3883 | -0.015 (-0.045, 0.015)<br>0.3152 |
|  | Some College or AA degree | -0.018 (-0.044, 0.008)<br>0.1843 | -0.018 (-0.044, 0.009)<br>0.1928 | -0.019 (-0.046, 0.008)<br>0.1753 | -0.021 (-0.049, 0.007)<br>0.1337 | -0.014 (-0.043, 0.015)<br>0.3374 |
|  | College Graduate or above | 0.005 (-0.021, 0.031)<br>0.7104 | -0.009 (-0.035, 0.018)<br>0.5180 | -0.003 (-0.030, 0.025)<br>0.8579 | 0.008 (-0.020, 0.037)<br>0.5643 | 0.017 (-0.010, 0.045)<br>0.2208 |
| SMOKE | Never | -0.020 (-0.047, 0.007)<br>0.1530 | -0.020 (-0.047, 0.007)<br>0.1445 | -0.029 (-0.057, -0.002)<br>0.0376 | -0.013 (-0.042, 0.016)<br>0.3885 | -0.019 (-0.048, 0.011)<br>0.2079 |
|  | Former | -0.008 (-0.033, 0.016)<br>0.5095 | -0.018 (-0.043, 0.006)<br>0.1370 | -0.010 (-0.035, 0.015)<br>0.4408 | -0.008 (-0.035, 0.019)<br>0.5690 | -0.001 (-0.028, 0.026)<br>0.9600 |
|  | Current | -0.028 (-0.045, -0.010)<br>0.0018 | -0.034 (-0.051, -0.016)<br>0.0002 | -0.028 (-0.046, -0.010)<br>0.0026 | -0.027 (-0.046, -0.008)<br>0.0057 | -0.025 (-0.044, -0.006)<br>0.0092 |
| ALP | Q1 | -0.024 (-0.047, -0.001)<br>0.0439 | -0.035 (-0.058, -0.011)<br>0.0037 | -0.033 (-0.056, -0.009)<br>0.0063 | -0.018 (-0.044, 0.007)<br>0.1577 | -0.014 (-0.039, 0.011)<br>0.2720 |
|  | Q2 | -0.003 (-0.029, 0.023)<br>0.8126 | -0.011 (-0.037, 0.016)<br>0.4388 | -0.003 (-0.030, 0.025)<br>0.8383 | 0.002 (-0.027, 0.030)<br>0.9060 | -0.002 (-0.031, 0.026)<br>0.8683 |
|  | Q3 | -0.016 (-0.043, 0.010)<br>0.2337 | -0.023 (-0.050, 0.003)<br>0.0852 | -0.012 (-0.040, 0.016)<br>0.4037 | -0.014 (-0.043, 0.015)<br>0.3326 | -0.019 (-0.048, 0.011)<br>0.2114 |
|  | Q4 | -0.044 (-0.069, -0.018)<br>0.0008 | -0.045 (-0.070, -0.019)<br>0.0006 | -0.049 (-0.076, -0.022)<br>0.0003 | -0.047 (-0.076, -0.019)<br>0.0010 | -0.036 (-0.064, -0.009)<br>0.0095 |

Age, sex, Had at least 12 alcohol drinks past one year?, Hypertension, Diabetes, Total 25(OH)D, BMI (obese, overweight, normal), TC(quartile groups), Ca (quartile groups), P (quartile groups), ALT (quartile groups), AST (quartile groups), CRP (quartile groups) were adjusted in the model

## Discussion

This study investigated the association between serum HDL-C and spinal BMD at different serum 25 (OH) D levels among Americans over 40 years old in a nationally representative sample. After adjusting all covariates, the weighted multiple linear regression model analysis showed a significant negative correlation between serum HDL-C and BMD in women aged 40-59 years with serum 25 (OH) D < 75nmol/L.

In the univariate analysis of serum HDL-C and spinal BMD, we found that there was a negative correlation between serum HDL-C and spinal BMD without adjustment of any covariable (Model1), sex, age and race (Model2) and after adjustment of all covariates (Model3)(Table 2). Some studies have found an inverse relationship between blood lipid levels and bone mass. For example, (1) adipocytes in lipid metabolism and osteoblasts in osteocytes are differentiated from MSCs(19); a long-term high-fat diet promotes MSCs to differentiate into adipocytes rather than osteoblasts, thus inhibiting bone formation. (2) High-fat environment promotes the transformation of osteoblasts into adipocytes in bone marrow(20). (3) High-fat environment enhances the bone resorption function of osteoclasts by increasing the levels of Type I collagen carboxy-terminal peptide (CTX-1) and Thrombospondin Related Adhesive Protein (TRAP)(21). (4) High-fat environment causes massive blood lipids and lipoproteins deposition in the arterial wall and the matrix of the subendothelial layer of bone vessels, resulting in lipid peroxidation. Lipid peroxides accelerated the inflammatory reaction of the arterial wall and inhibited the differentiation and bone mineralization of osteocytes(22). This significant relationship is indistinguishable and includes all the components tested in the blood fat spectrum: TC, HDL-C, Low-density lipoprotein cholesterol (LDL-C) and Triglycerides (TG)(23). However, few studies on the correlation and mechanism between increased HDL-C and decreased BMD. Kha et al. reported that specific oxysterol could stimulate MSCs to differentiate into osteoblasts, while high serum HDL-C level can remove oxysterol from peripheral tissue, which has a negative impact on osteogenic differentiation(24). Dennison et al. found that serum high serum HDL-C level is related to higher fat content and higher body weight. There is a negative correlation between BMD and serum HDL-C in women, but the correlation decreases after adjusting the fat ratio(25). Jirapinyo et al. observed that oral estrogen/progesterone combination increased BMD but decreased serum HDL-C levels in postmenopausal women(26). Mazidi et al. found that serum HDL-C was positively correlated with inflammatory markers such as C-reactive protein, leukocyte and fibrinogen(27). Therefore, it can be speculated that serum HDL-C and bone metabolism mechanisms may be related to hormone deficiency and inflammatory response.

In addition, the correlation between serum HDL-C and spinal BMD was different at separate serum 25 (OH) D levels. Serum HDL-C was a negative correlation with total spine and L1-L4 BMD at serum 25 (OH) D < 75nmol/l(P<0.05). However, when serum 25 (OH) D ≥ 75nmol/l, there was no correlation between total spine, L1-L4 BMD and HDL-C(P>0.05)(Table 3). One possible mechanism is that 1,25-(OH) 2D3 in serum 25 (OH) D can inhibit the secretion and mRNA expression of apolipoprotein AI (the main apolipoprotein of HDL-C), which makes the activity of apolipoprotein AI promoter decrease, affects the level and function of HDL-C(28-30), and then weakens the process of bone regeneration inhibition in the high-fat environment. Another possible mechanism is a positive correlation between serum 25 (OH) D and BMD(28, 31), which reduces or even eliminates the correlation between serum HDL-C and BMD. The molecular mechanism is that serum 25 (OH) D regulates calcium homeostasis by affecting intestinal calcium absorption, renal calcium reabsorption and osteoclast bone resorption(32). Serum vitamin D can act on vitamin D response elements of osteoblasts and bone nuclei, and regulate the expression of osteocalcin, low-density lipoprotein receptor-related protein 5, fibroblast growth factor 23, type I, collagen and other proteins, which has a substantial regulatory effect on bone turnover and bone mineralization(33).

To further explore the correlation between BMD and serum HDL-C and its possible mechanism in different populations with serum 25 (OH) D < 75nmol/L, we carried out a stratified analysis of sex and age in all populations with serum 25 (OH) D < 75nmol/L. The results showed that the correlation between serum HDL-C and BMD varies with gender and age when serum 25 (OH) D < 75 nmol/L. The negative correlation between serum HDL-C and BMD only exists in the female population, especially those 40-59 years. It is similar to previous studies that have shown that the serum HDL-C level in postmenopausal women with osteopenia or OP is higher(17, 34, 35). There are several possible reasons for this difference. First, there are differences in the pathogenesis of osteoporosis between men and women. Hypercalciuria, abnormal testosterone metabolism level, hormone disorder related to bone metabolism and hypogonadism caused by aging are the leading causes of osteoporosis in older men(36), while estrogen deficiency caused by drastic changes of sex hormone levels in women aged 40-59 years may be the leading cause of OP in this age group(17, 37). Estrogen deficiency can increase apoptosis of osteoblasts, which can act on the estrogen receptorα(ERα) target of osteoblasts to inhibit the differentiation of osteoblasts and increase apoptosis, and also impede the production of receptor activator of nuclear factor-κB (RANK) by osteoblasts, T cells and B cells, and reduce the activity of osteoblasts (38). Estrogen deficiency in women before and after menopause will lead to a decrease in the level of osteoprotegerin (OPG), promote local bone inflammation and promote the expression of inflammatory cytokines. The increased expression of inflammatory factors can also enhance the activity of osteoclasts, which further leads to the aggravation of bone mass loss(39). Estrogen deficiency breaks the balance between bone formation and bone resorption(40). This process is not significant in elderly male. Secondly, studies have shown that changes in menopause can lead to distraction and depression, while depression may increase the risk of osteoporosis(41). Women’s overall level of physical activity is lower than that of men(42). The gender differences in bone size and geometry, bone histomorphometry and sex hormones may be the reasons for the difference in bone mass and degree of osteoporosis between men and women(43).

Most studies only confirmed a negative correlation between serum HDL-C and BMD. This study investigated the relationship between serum HDL-C and BMD by stratified analysis of sex, age, serum 25 (OH) D level, and all covariables. The results showed a significant negative correlation between serum HDL-C and BMD in women aged 40-59 years old with serum 25 (OH) D < 75nmol/L. This correlation did not exist in women over 60 years old with serum 25 (OH) D < 75nmol/L, all males and all populations with 25 (OH) D ≥ 75nmol/L. Our results are more accurate and specific than the previous simple negative correlation between serum HDL-C and BMD. This was of great significance for clinicians and this population to closely observe BMD and early intervention to prevent OP. In addition, this study based on NHANES database data, the sample size is large, the clinical data are sufficient, and the study population based on the general population, which may be more representative than the population recruited from the hospital.

In this study, we also found that the β value and significance of the correlation between L1-L4BMD with serum HDL-C decreased in turn (Table 2), which indicated that the L1 BMD is significantly affected by the increase of serum HDL-C level, and the correlation was the most significant, followed by L2, L3, L4. The comparison of L1-L4 BMD in the original data of the included population also found that the relationship between lumbar BMD size of all subjects in the population was L1< L2 < L3 < L4 (Fig 3). A reasonable explanation may be that the decrease of spinal BMD begins at L1 or L1 BMD is more affected by bone mass loss caused by various factors such as aging, and this relationship was also found in a census study of lumbar vertebrae BMD(44).

**Figure3.**
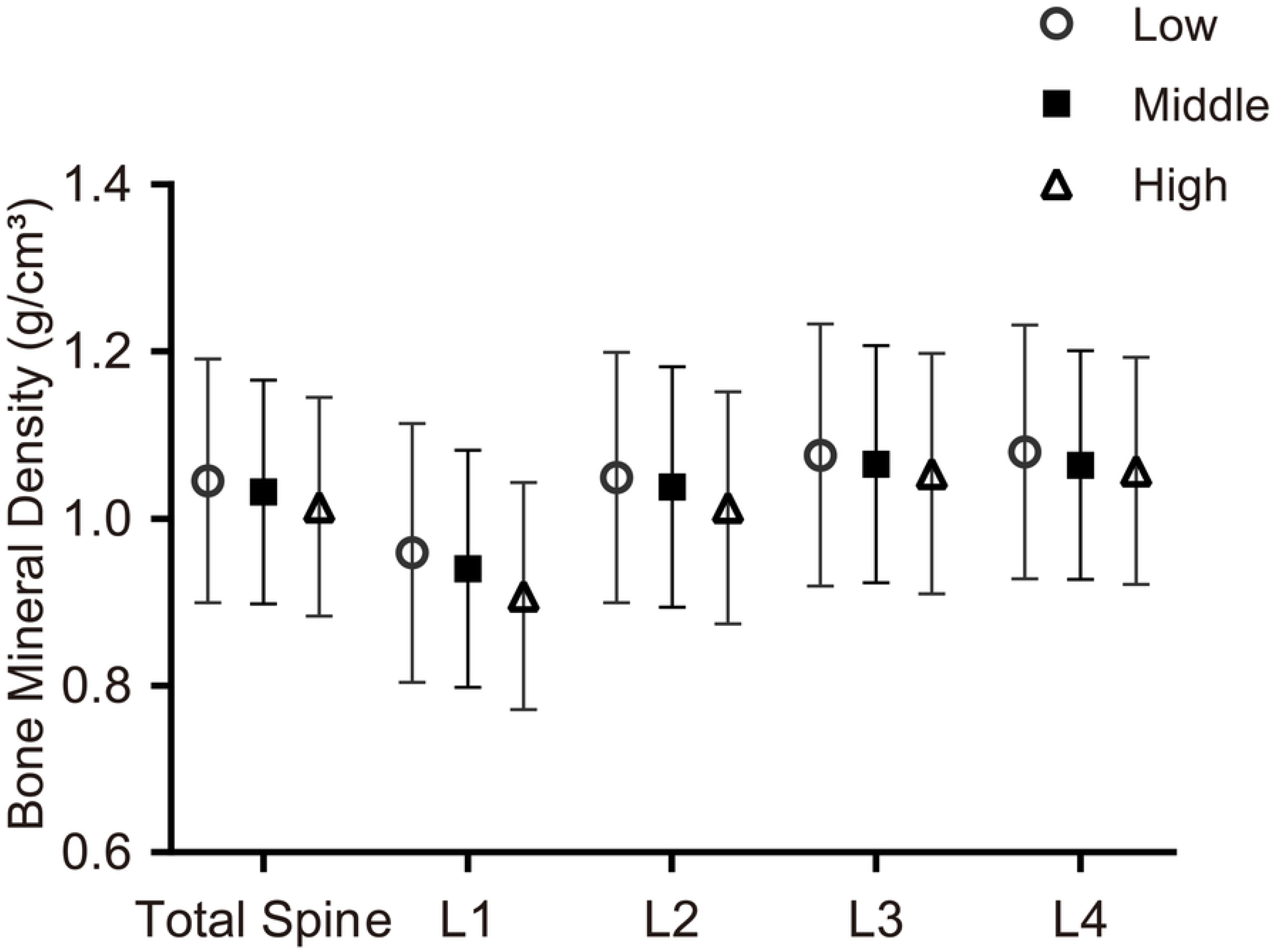

The limitations of the study are stated below:

1. Because of its cross-sectional design, this study cannot accurately detect the causal relationship between serum HDL-C, serum 25 (OH) D levels and BMD.
2. We excluded patients who had kidney disease, treated osteoporosis, and took prednisone or cortisone tablets almost every day, so this study’s conclusions could not be applied to them.
3. Collecting questionnaire data through questionnaires and interviews may lead to recall bias and affect the study’s conclusions.
4. All the participants in this study are American residents, and the conclusions may not apply to all populations and races.

In general, more extensive data, rigorous experimental and multi-center designs are needed in the future to verify further the conclusion that serum HDL-C is negatively correlated with BMD, as well as more fundamental studies on the molecular level of HDL-C and bone metabolism to obtain a relatively more objective and reliable evidence-based basis.

## Data Availability

All files are available from the NHANES database.

https://wwwn.cdc.gov/nchs/nhanes/Default.aspx

## Supporting information

**S1 Fig**. Flow chart of participants selection.

**S2 Fig**. Classification of Women aged 40-60 years with serum 25(OH)D<75nmol/l based on the tertile interval of HDL-C concentration, and the association between different HDL-C groups and BMD.

**S3 Fig**. Association between BMD and L1-L4 of adults over 40 years old.

**S1 Table. Characteristics of study sample: US population aged ≥40 years in the NHANES 2007-2010**. NHANES = National Health and Nutrition Examination Survey; SD = standard deviation; Values are mean ± SD or n (%).HDL-C = high-density lipoprotein cholesterol; BMI = Body mass index; CRP = C-reactive protein; TC = Cholesterol; ALP = Alkaline phosphatase; ALT = Alanine aminotransferase; AST = Aspartate aminotransferase; BMD = bone mineral density; CI = confidence interval; β regression coefficient

**S2 Table. Correlation between high-density lipoprotein-cholesterol and bone mineral density**. Model1, no covariates were adjusted Model2, age,sex,race / ethnicity, were adjusted Model3, age, sex, race/ethnicity, Education, Smoking, Had at least 12 alcohol drinks past one year?, Marital status, Hypertension, Diabetes, Total 25(OH)D, BMI (obese, overweight, normal), TC (quartile groups), Ca (quartile groups), P (quartile groups), ALP (quartile groups), ALT (quartile groups), AST (quartile groups), CRP (quartile groups) were adjusted in the model.

**S3 Table. Correlation between high-density lipoprotein cholesterol and bone mineral density based on 25(OH)D status classification**. Age, sex, race/ethnicity, Education, Smoking, Had at least 12 alcohol drinks past one year?, Marital status, Hypertension, Diabetes, BMI (obese, overweight, normal), TC (quartile groups), Ca (quartile groups), P (quartile groups), ALP (quartile groups), ALT (quartile groups), AST (quartile groups), CRP (quartile groups) were adjusted in the model.

**S4 Table. Correlation between high-density lipoprotein cholesterol and bone mineral density based on Gender and Age status classification**. Race/ethnicity, Education, Smoking, Had at least 12 alcohol drinks past one year?, Marital status, Hypertension, Diabetes, Total 25(OH)D, BMI (obese, overweight, normal), TC (quartile groups), Ca (quartile groups), P (quartile groups), ALP (quartile groups), ALT (quartile groups), AST (quartile groups), CRP (quartile groups) were adjusted in the model.

**S5 Table. Correlation between high-density lipoprotein cholesterol and bone mineral density based on covariate status classification**. Age, sex, Had at least 12 alcohol drinks past one year?, Hypertension, Diabetes, Total 25(OH)D, BMI (obese, overweight, normal), TC(quartile groups), Ca (quartile groups), P (quartile groups), ALT (quartile groups), AST (quartile groups), CRP (quartile groups) were adjusted in the model

**List of Abbreviations**

BMD, bone mineral density; HDL-C, high-density lipoprotein cholesterol; 25(OH)D, 25-hydroxyvitamin D; OP, Osteoporosis; NHANES, National Health and Nutrition Examination Survey; SD, standard deviation; CRP, C-reactive protein; BMI, body mass index; TC, Cholesterol; ALP, Alkaline phosphatase; ALT, Alanine aminotransferase; AST, Aspartate aminotransferase; CI, confidence interval

## Acknowledgements

None

## Author Contributions

**Conceptualization:** Peng Gu, Bin Pu, WeiDong Luo.

**Data Curation:** BaiHang Chen.

**Formal Analysis:** Peng Gu, Bin Pu, XiaoHui Zheng, ZhanPeng Zeng, WeiDong Luo.

**Investigation :** Peng Gu, Bin Pu, BaiHang Chen.

**Methodology:** BaiHang Chen.

**Project Administration:** WeiDong Luo.

**Supervision:** WeiDong Luo.

**Validation:** XiaoHui Zheng, ZhanPeng Zeng.

**Visualization :** BaiHang Chen.

**Writing – Original Draft Preparation:** Peng Gu, Bin Pu, WeiDong Luo.

**Writing – Review & Editing:** Peng Gu, Bin Pu, WeiDong Luo.

## Ethics approval and consent to participate

The original survey was approved by the NCHS Research Ethics Review Board and all adult participants provided written informed consent. The present analysis was deemed exempt by the Institutional Review Board at our institution, as the dataset used in the analysis was completely de-identified.

## Data availability

Some or all data generated or analyzed during this study are included in this published article or in the data repositories listed in References.NHANES data is available publically at https://wwwn.cdc.gov/nchs/nhanes/Default.aspx

